# Open educational resources on how to conduct randomised clinical trials: a protocol for a landscape analysis

**DOI:** 10.1101/2024.02.16.24302873

**Authors:** Kim Boesen, Sarah Louise Klingenberg, Christian Gluud

**Affiliations:** Copenhagen Trial Unit, Centre for Clinical Intervention Research, The Capital Region, Copenhagen University Hospital ─ Rigshospitalet, Copenhagen, Denmark; Department of Regional Health Research, The Faculty of Health Sciences, University of Southern Denmark, Odense, Denmark

## Abstract

This is a protocol for the project entitled, “Open educational resources on how to conduct randomised clinical trials”.

## Introduction

A large part of conducted randomised clinical trials across fields, indications, and healthcare interventions are not informative for clinical practice and is considered waste.^1-8^ The three main reasons for clinical trials being uninformative are that they are done for the wrong reason, they are designed the wrong way, and/or their results are not analysed and reported adequately. ^1-8^ As Doug Altman put it in his famous essay on the scandal of poor research from 1994, “We need less research, better research, and research done for the right reasons”.^9^

For decades, systematic reviews and assessments of clinical trials across many different fields and specialties have reported issues with risk of bias, suboptimal trial designs, underreporting, selective outcome reporting, and publication bias, colloquially referred to as ‘research waste’.^1-8^ Recently, assessments of clinical trials’ informativeness encompassing indicators also of feasibility, transparency, and reproducibility, have emerged. One study of 125 diabetes, ischaemic heart disease, and lung cancer trials reported that only 26% fulfilled the authors’ four criteria of informativeness (clinical question, design, feasibility, reporting).^10^ Another similar study of 347 gynecology and obstetrics trials reported that only 10% fulfilled minimum half of the authors’ eight prespecified ‘usefulness’ criteria (clinical question, preceding systematic review, power, pragmatic, patient oriented, financially worth it, transparency).^11^ One main reason for this lack of ‘informativeness’ may be that it is not a priority during medical, graduate, and postgraduate training to learn about proper conduct of clinical trials, and access to freely available quality educational material may also be sparse.

We want to make a landscape analysis of publicly available online educational material on how to conduct interventional clinical trials, covering all aspects from the initial ideation to the design, conduct, and final reporting. Rather than focusing on traditional book or paper-based material, we will focus on free online material, colloquially known as open educational resources (OER),^12, 13^ and primarily in the ‘microlearning’ format,^14, 15^ i.e. short (or ‘bite sized’) concise videos, tutorials, or instructions. Such material is often used in a ‘blended learning’ setup combining online and face-to-face classes with video and written material and integrating various technologies.^16-19^ Within some medical specialties, particularly anaesthesiology and critical care medicine, extensive online communities with freely available educational material exist, known as free open access medical education (FOAM).^20-22^ Overviews and surveys of OER in various specialties, including pediatric intensive care,^23^ nephrology,^24^ radiation oncology,^25^ anaesthesiology,^26^ pediatrics,^27^ critical care,^28, 29^ otolaryngology,^30^ and medical physics^31^ report many available resources, including videos, podcasts, and tutorials. We will use this landscape analysis as a primer to build a comprehensive blended learning curriculum on randomised clinical trial conduct targeting clinicians, investigators, trialists, researchers, and anyone interested in the topic on how to make fair tests of treatments.^32, 33^

## Research objectives

1. To identify publicly available educational material or open educational resources (videos and/or written material) about the conduct of randomised clinical trials.
2. To describe and categorise existing material:
  a. Curriculum, i.e. what content is covered?
  b. Detail level, i.e. how thorough and comprehensive is the content?
  c. Documentation, i.e. is the material evidence based and reference based?
  d. Format, i.e. is the material written, videos, blended combination of video and written, or other combinations, and are there interactive elements, such a quizzes or questionnaire tests?

## Methods

Due to the unconventional topic, we will do (i) a conventional systematic search of databases of published literature, and (ii) we will also search other (unconventional) resources that one could expect researchers and clinicians to look for open educational resources on randomised clinical trials, such as YouTube, Google, and dedicated online learning portals.

### Eligibility criteria

We search for any freely available, instantaneously accessible, online course or educational resource, or massive open online courses (MOOC) as they may be referred to, covering any aspect of interventional clinical trial conduct. We do not apply restrictions on language, publication year, extent, and content.

#### (i) Conventional database searches

Our information specialist (SLK) will search MEDLINE ALL Ovid, Embase Ovid, Science Citation Index Expanded (Web of Science), Social Sciences Citation Index (Web of Science), Conference Proceedings Citation Index – Science (Web of Science), Conference Proceedings Citation Index – Social Science & Humanities (Web of Science), and Cumulative Index to Nursing and Applied Health Literature (CINAHL; Ebsco Host) using a compound search string consisting of keywords relevant to the field (Appendix 1).

We will merge and import the combined hits into the open-source artificial intelligence (AI) assisted screening program for systematic reviews, ASReview.^34^ One author (KB) will screen the hits in a one-step process (i.e. title/abstract and, when needed, full-text) to make an immediate final decision about inclusion or exclusion. We will apply an arbitrary stopping rule of 100 consecutive irrelevant hits, i.e. meaning that we will not necessarily screen all retrieved hits, but instead we will screen only those hits deemed most relevant by ASReview.^35^

For the ASReview machine learning algorithm to be able to rank and order the full number of retrieved hits, it must be trained (i.e. supervised learning) on a minimum of two relevant references (in our case, publications describing open educational resources on randomised clinical trials), and two irrelevant references. At our current stage, we are not aware of relevant references. One solution would be to train the algorithm on overview articles pertaining to OER in specific medical fields^20-31^ or to train it on articles describing specific OERs, such as online residency training programs.^36-38^ As a last resort, we can train it on those articles and screen all retrieved articles. However, this may not be feasible if the systematic database search returns a very large number of hits (e.g. >10.000 hits). We will decide our final strategy based on the number of retrieved hits and if we discover more eligible publications before screening.

#### (ii) Searching other resources

We will use a two-step search strategy by first doing ‘cold searches’ of general websites using expected key words to identify ‘seed references’ such as relevant material, videos, and resources. One author (KB) will use these keywords and terminology to subsequently stringently search the listed resources and others, if needed.

##### General website searches

We will search common websites to find seed videos and courses to guide our search, including YouTube (https://www.youtube.com/), Google (https://www.google.com/), and Bing (https://www.bing.com/). An example of a search string: “online learning material clinical trials”.

We know of one comprehensive resource already: the YouTube channel ‘GCP-Mindset’.^39^ The channel is maintained by the contract research organisation company GCP-Service International^40^ and they have published 350 videos (by 1 February 2024; not all relevant) on matters related to clinical trial conduct.

##### Online learning portals

We will search the most common and used online learning portals, including:

1. **Coursera** (https://www.coursera.org/) is one of the most popular and largest online learning portals with more than 5.800 courses. It was founded in 2012 by Stanford University but is now a for-profit company.
2. **EdX** (https://www.edx.org/) has more than 4.000 courses available. It was founded by Harvard University and Massachusetts Institute of Technology (MIT) in USA in 2012 but it is now a subsidiary of the for-profit company 2U.

Portals with paid only material, such as Udemy (https://www.udemy.com/), or restricted access, such as Udacity (https://www.udacity.com/), will not be considered.

##### University portals

We will also search individual major US and European university eLearning portals. There might be overlap with the courses provided on Coursera and EdX.

1. Stanford Online (https://online.stanford.edu/)
2. Harvard Professional and Lifelong learning (https://pll.harvard.edu/)
3. MIT Open course ware (https://ocw.mit.edu/)
4. SwissMOOC (https://www.swissmooc.ch/), is a collaboration of most Swiss universities’ online courses.
5. Oxford University Online and distance courses (https://www.ox.ac.uk/admissions/conBnuing-education/online-and-distance-courses).
6. Cambridge Advance Online (https://advanceonline.cam.ac.uk/)

##### Clinical trial units

We will map major Scandinavian, European, and international clinical trial units and clinical trial networks for relevant educational material. The following list may not be exhaustive:

1. European Clinical Research Infrastructure Network (ECRIN) (https://ecrin.org/), and the national charters.
2. Swiss Clinical Trial Unit Network (https://www.scto.ch/en.html), including individual CTUs, e.g. CTU Bern (https://www.ctu.unibe.ch/) and University of Zurich Clinical Trials Center (https://www.usz.ch/en/clinic/clinical-trials-center/).
3. Danish trial units, e.g. the Open Patient data Explorative Network at Southern University Denmark (https://open.rsyd.dk/), Aarhus University’s Clinical Trial Unit (https://clin.au.dk/research/clinical-trial-unit), and the centralised Good Clinical Practice (GCP) Unit (https://gcp-enhed.dk/).
4. Clinical Studies Sweden (https://kliniskastudier.se/) and it’s six regional websites.
5. Clinical Trials Unit at Oslo University Hospital (https://forskerstoVe.no/en/kliniske-studier) and Clinical Research Unit at region Mid-Norway (https://www.klinforsk.no/).
6. Medical Research Council Clinical Trials Unit (https://www.mrcctu.ucl.ac.uk/).
7. University of Oxford Clinical Trial Units (https://www.ctsu.ox.ac.uk/research).
8. Cambridge Clinical Trial Units (https://cctu.org.uk/).
9. Stanford Clinical and Translational Research Unit (https://med.stanford.edu/ctru.html).

##### Other online resources for clinical research conduct

If, and when, we identify other resources containing educational resources on the conduct of interventional clinical trials that do not fit the first four categories, they will be listed here.

Examples of such resources include:

1. The UK Faculty of Public Health’s Health Knowledge portal (https://www.healthknowledge.org.uk/).
2. The European Communication on Research Awareness Need (ECRAN) Project (http://ecranproject.eu/en), which is a European Commission project on communicating the importance of clinical trials.

## Discussion

The published literature on so called ‘microlearning’ is growing,^12, 13^ and even more so is the literature on open educational resources (OER) and free open access medical education (FOAM) across medical specialties.^20-31^ Even the concept of peer reviewed medical education videos has emerged.^41^ However, based on basic preliminary searches (Appendix 2) we expect some, but not many, relevant online resources on how to plan, launch, execute, analyse, and report randomised clinical trials. We expect videos and courses to be rather high-level, such as focusing on good clinical practice (e.g. the YouTube ‘GCP-Mindset’ channel^34^), whereas educational material focusing on the individual steps necessary to conduct a rigorous, reproducible, and reliable clinical trial may be scarce.

Material made available on Coursera and EdX are mainly produced by prestigious US universities, e.g. Johns Hopkins or Stanford, and this needs to be taken into consideration when evaluating the content and its global appeal and usefulness. Especially if the content focuses on local or regional legislation, it may not be applicable in other regions. Another more important limitation to open educational resources may be a lack of scientific rigour and quality, and content may be more or less evidence based.^21, 42, 43^ However, we do not intend to make a detailed qualitative assessment of the identified resources, which will be beyond the scope of this landscape analysis.

As a minimum we will make our full results available on the Copenhagen Trial Unit’s web site (www.ctu.dk) as part of our forthcoming blended learning curriculum. If we judge the results to have appeal for a larger audience, we will also seek publication in a regular journal, to put our findings into a wider context.

## Data Availability

The complete dataset will be made publicly available alongside the final report.

## Acknowledgements

KB wrote the first draft to this protocol and KB, SLK, and CG revised it, and all agreed on the final version.

## Conflicts of interest

KB and CG will develop a publicly available, freely accessible blended learning curriculum on randomised clinical trials. They have no financial or otherwise intellectual conflicts related to the topic.

## Data sharing statement

The complete dataset will be made publicly available alongside the final report.

## Funding statement

This project does not receive any funding.

## Appendix 1

### MEDLINE ALL Ovid (1946 to the date of the search)

1. (‘open access’ adj3 (education* or web-site* or meducation*)).ti,ab.
2. (((online or video or digital or web-based) adj3 (resource* or material* or program* or education* or training* or course* or learning*)) or e-learning or ‘blended learning’ or ‘micro learning’).ti,ab.
3. 1 or 2

### Embase Ovid (1974 to the date of the search)

1. (‘open access’ adj3 (education* or web-site* or meducation*)).ti,ab.
2. (((online or video or digital or web-based) adj3 (resource* or material* or program* or education* or training* or course* or learning*)) or e-learning or ‘blended learning’ or ‘micro learning’).ti,ab.
3. 1 or 2

### CINAHL (Ebsco host; the date of the search will be given at review stage)

S3 S1 OR S2

S2 TX ((((online or video or digital or web-based) N3 (resource* or material* or program* or education* or training* or course* or learning*)) or e-learning or ‘blended learning’ or ‘micro learning’)) OR AB ((((online or video or digital or web-based) N3 (resource* or material* or program* or education* or training* or course* or learning*)) or e-learning or ‘blended learning’ or ‘micro learning’))

S1 TX ((‘open access’ N3 (education* or web-site* or meducation*))) OR AB ((‘open access’ N3 (education* or web-site* or meducation*)))

**Science Cita3on Index Expanded (1900 to the date of the search), Social Sciences Cita3on Index (1956 to the date of the search), Conference Proceedings Cita3on Index – Science (1990 to the date of the search), Conference Proceedings Cita3on Index – Social Science & Humani3es (1990 to the date of the search) (Web of Science)**

#3 #2 OR #1

#2 TI=(((online or video or digital or web-based) N3 (resource* or material* or program* or education* or training* or course* or learning*)) or e-learning or ‘blended learning’ or ‘micro learning’) OR AB=(((online or video or digital or web-based) N3 (resource* or material* or program* or education* or training* or course* or learning*)) or e-learning or ‘blended learning’ or ‘micro learning’)

#1 TI=(‘open access’ N3 (education* or web-site* or meducation*)) OR AB=(‘open access’ N3 (education* or web-site* or meducation*))

## Appendix 2

Initial pilot searches

### Coursera

Search words: “Clinical trials” Relevant courses:

1. Johns Hopkins University - ‘Design and Interpretation of Clinical Trials’ https://www.coursera.org/learn/clinical-trials Six modules, rather short course. Voiced-over PowerPoint slides, no interactive elements or background material readily available.
2. Johns Hopkins University - “Design and conduct of clinical trials” https://www.coursera.org/learn/design-and-conduct-clinical-trials. Five modules, similar format.

### EdX

Search words: “Clinical trials”

Relevant courses:

1. Stanford University - Thinking critically (https://www.edx.org/learn/critical-thinking-skills/stanford-university-thinking-critically-interpreting-randomized-clinical-trials). Short overview of the topic, not about the whole process of making a clinical trial.
2. MD Anderson - Certain Pragmatic trials (https://www.edx.org/learn/research/the-university-of-texas-md-anderson-cancer-center-in-houston-certain-pragmatic-clinical-trials-and-healthcare-delivery-evaluations).

Short overview of the topic, not about the whole process of making a clinical trial.

